# Predicting Drug-Resistant Tuberculosis Treatment Outcomes: A Leakage-Controlled Evaluation of Topological Data Analysis Augmentation

**DOI:** 10.64898/2026.10.23.26366286

**Authors:** Darlington Tozivepi Mutakwa, Maria Vivien Visaya, Faraimunashe Chirove

## Abstract

**Background:** We asked what topological structure adds to a clinical prediction model and how that addition should be measured. Drug-resistant tuberculosis (DR-TB) treatment outcome prediction is the testbed. The contribution is an evaluation design and a bounded null result.

**Methods:** Cross-sectional records for 9210 patients from the NIAID TB Portals programme were collated to one row per patient by worst-case aggregation, giving a 99-feature clinical schema. We derived two topological representations. A Mapper graph, built on a UMAP lens over a Gower dissimilarity, supplied four neighbourhood-position features. Local Vietoris–Rips persistent homology on each patient’s 26-point neighbourhood supplied degree-0 and degree-1 summaries, vectorised by the best of thirteen catalogued methods. We compared five feature configurations across four classifiers in a pipeline in which every topological step is fitted inside the training partition. A sealed test lockbox of 1776 patients, 463 of them with adverse outcomes, was opened once. We compared discrimination by the area under the receiver operating characteristic curve (AUROC) using paired DeLong tests with Bonferroni correction (corrected threshold 0.0125).

**Results:** No topologically augmented configuration meaningfully improved lockbox AUROC over the clinical-only baseline under any classifier. The best AUROC was 0.7548, achieved by the baseline with XGBoost. Across all sixteen comparisons the upper 95% confidence limit never exceeded **+0.014** AUROC. Five comparisons met the corrected threshold. Four favoured the baseline, and one, under the support vector machine, favoured the augmented model by a negligible **+0.0005** AUROC. The same features supported neighbourhood-based risk stratification and revealed variation in adverse-outcome composition across patient neighbourhoods: adverse-outcome purity ranged from 5.9% to 30.5% across the fifteen largest Mapper nodes, stably under resampling.

**Conclusions:** The findings support a complementary framing of topological data analysis in clinical prediction. Topological features are best understood not as replacements for clinical predictors but as an analytical lens that reveals geometric structure in the patient population not captured by the clinical feature representation alone.

## 1 Background

Drug-resistant tuberculosis (DR-TB) continues to cause significant morbidity and mortality worldwide. The 2025 WHO Global Tuberculosis Report estimates that around 400,000 people develop multidrug-resistant or rifampicin-resistant tuberculosis annually, with global treatment success rates remaining below 60% [1]. Identifying patients at high risk of treatment failure from their clinical record would allow earlier escalation of care and more efficient allocation of resources, but prediction from baseline clinical data is constrained. Models built on demographic, clinical and radiological variables typically report AUROC values in the range of 0.71 to 0.74 for treatment failure and loss to follow-up [2]. Multimodal analyses incorporating genomic and imaging data have improved discrimination only for narrow endpoints such as culture conversion at six months [3]. Prediction of TB treatment failure from any single data modality plateaus at a level that is promising but insufficient, and meaningful gains in discrimination require integrating clinical, radiological, and pathogen genomic features into a unified multimodal model [4].

Topological data analysis (TDA) offers a different perspective on patient data. Rather than summarising each patient as a flat feature vector, TDA constructs representations of the geometric shape of the data distribution. TDA investigates which patients cluster together, where the boundaries between groups lie, and whether the population exhibits structural patterns that a coordinate-based representation would not capture. The premise is that this geometric information is not already contained in standard predictors and that recovering it could improve prediction accuracy. If topological features carry discriminative signal beyond a well-specified clinical baseline, augmenting that baseline should improve AUROC on an independent test set. That is the specific claim this paper evaluates.

Applied studies of TDA in clinical prediction have rarely tested this claim carefully. Three problems recur. First, few studies specify the clinical baseline rigorously, so the comparison is between a topological model and a weakly configured reference rather than between topological augmentation and a well-tuned incumbent [5]. Second, topological features such as Mapper nodes and persistence diagrams are computed from pairwise dissimilarities or embeddings fitted on the full cohort, so they can carry information from held-out patients into the training representation, a form of leakage documented as a recurring failure across machine learning research [6]. Third, differences in AUROC are typically reported as point estimates without a paired test or a multiplicity correction, so a positive result cannot be distinguished from sampling variation and a null result cannot be bounded. Together these problems mean that neither positive nor negative results in this literature are as informative as they appear.

This paper makes three contributions, each bearing on how TDA augmentation should be evaluated. First, we describe a leakage-controlled evaluation design in which every topological step — the dissimilarity matrix, the UMAP embedding, the Mapper cover and the local persistence computation — is fitted exclusively on the training partition and applied to a sealed test set without further modification. Second, we report a bounded null result: no topologically augmented configuration meaningfully improved discrimination over a 99-feature clinical baseline on a held-out lockbox of 1,776 patients, with all upper confidence limits below +0.014 AUROC across sixteen paired DeLong tests with Bonferroni correction. Third, we show that this null is compatible with genuine structural information in the topological features, information that a cohort-level rank statistic is not designed to detect, and we propose a minimal set of evaluations capable of detecting it. The DR-TB setting makes this a demanding testbed: the baseline is already well-specified, and any incremental contribution must be measured against it.

### 1.1 Related work

#### 1.1.1 Topological data analysis in clinical prediction

Topological data analysis (TDA) has been applied across biomedicine for over a decade and a recent review of that work has been done by Skaf and Laubenbacher [7]. The claims in it are of two kinds and separating them matters. The first is a discovery claim. Nicolau and colleagues [8] analysed breast cancer gene expression topologically and isolated a subgroup with an unusual survival profile that conventional stratification had missed. Li and colleagues [9] built a patient similarity network from electronic health records and genotype data and reported three subtypes of type 2 diabetes with distinct comorbidity profiles. Since 2015, TDA has identified discriminative structure across multiple biomedical domains. In neurotrauma, TDA identified perioperative hypertension as a superior predictor of recovery [10]. In neuroscience, clique topology revealed that hippocampal neural correlations encode intrinsic circuit geometry beyond spatial position [11]. For epilepsy, a persistent-entropy classifier distinguished seizure from healthy electroencephalography with 97.2% AUROC [12]. In histology, persistent entropy differentiated epithelial tissue types [13], while in oncology, TDA characterised dynamic tumour vascular network structure in relation to treatment response [14]. None of these studies asserts that a topological feature improves the discrimination of a tuned prediction model, so none is put at risk by a study testing only that narrower proposition. The second is a performance claim. Kazeminia et al. [15] showed that topological descriptors embedded within Multiple Instance Learning architectures yielded AUROC gains of 3.3% to 5.9% over standard pooling strategies.

Incorporating persistent homology features into convolutional and vision-transformer architectures for breast ultrasound screening similarly yielded consistent improvements in both accuracy and AUC across multiple backbone architectures, even under limited data conditions [16]. In another context, a topology-guided network achieved a 4.6% improvement in AUROC for predicting neoadjuvant chemotherapy response [17]. These results indicate that topological features can improve predictive performance when applied to imaging data, where topology is computed directly from pixel-level structure or from learned embeddings. The dominant condition appears to be that topological information contributes most when the underlying feature representation is noisy or weakly discriminative and adds marginal benefit once embeddings are already well-structured [15]. This raises the question of whether such gains generalise to settings where the baseline feature space is already highly informative, such as curated clinical tabular data.

#### 1.1.2 Mapper and persistent homology as feature generators

Two constructions dominate the TDA space. Mapper takes a dissimilarity on the observations and a filter function, covers the filter range with overlapping intervals, clusters the observations in each preimage and forms a graph with one node per cluster and an edge wherever two clusters share an observation [18]. Persistent homology takes a filtration, a nested family of simplicial complexes indexed by a scale parameter, and records the scale at which each connected component, loop or higher void appears and disappears. Its output is a persistence diagram, a multiset of birth and death pairs [19]. Both outputs are objects, not vectors, and most clinical applications simply inspect them as is. However, using them as predictors requires vectorisation, a map to a fixed-length real vector. For persistence diagrams, this is done with landscapes [20], persistence images [21], kernels [22] and a wider family catalogued elsewhere [23]. For Mapper, the usual route is graph descriptors attached to each observation through the nodes containing it.

Vectorising a topological object produces a fixed-length feature block subject to the same requirements as any other predictor set: its dimension must be proportional to the available data, it may correlate with existing features, and fitting the vectorisation on the full cohort before splitting introduces leakage. Stability results establish that persistence diagrams change by a bounded amount under small perturbations of the input [24]. Neither result establishes whether a vectorised topological summary adds information beyond what the original feature set already contains. That is an empirical question, and one this paper addresses directly.

#### 1.1.3 Mixed-type clinical data and the dissimilarity problem

Every topological pipeline sits downstream of a dissimilarity. For imaging and omics data this is rarely contentious. Tabular clinical data are different. A patient record mixes continuous measurements, ordered severity grades, unordered categories, binary indicators, and no canonical dissimilarity exists for that mixture.

Gower’s coefficient is the standard answer, averaging a per-variable dissimilarity across the record [25]. It also fixes, by construction rather than by evidence, the weight of a single category disagreement against a full-range difference in a continuous variable. Van de Velden and colleagues review the wider family of mixed-data distances and show the choice changes recovered cluster structure materially [26]. The consequence for the topology is direct. Stability theorems control the output under small perturbations of a fixed metric, so they say nothing about two metrics that are not close to each other. A Mapper graph on mixed clinical data is therefore partly a property of the encoding, as is any persistence computation built on the same dissimilarity or on an embedding derived from it [27]. Clinical applications typically name the metric in one clause and report no sensitivity analysis.

#### 1.1.4 Gaps in current evaluation practice

The weakness of this literature is not in the topology but in the comparison. We identify four problems. The first is the baseline. An incremental claim needs a reference model tuned as carefully as the augmented one. Many topological studies report the topological model alone, or compare against a default configuration of a conventional model, so the claim that topology improves prediction has no denominator. The general form of the point is settled: a systematic review found no benefit of machine learning over logistic regression once baselines were properly specified [5]. The second is leakage. Topological features are computed from the geometry of a set of observations, which makes them structurally prone to it. A dissimilarity matrix, a Mapper cover or an embedding fitted on the full cohort before splitting carries information about held-out observations into the training representation. Kapoor and Narayanan document leakage as a recurring failure across seventeen fields and 294 papers [6]. The topological case is a clean instance of it and is rarely addressed explicitly. The third is inference. Differences in AUROC are usually reported as point estimates. A paired test for correlated ROC curves has been available since 1988 [28], and comparing several feature sets across several classifiers also requires a multiplicity correction. Without both, a difference of 0.01 supports no conclusion in either direction. The fourth is the choice of metric. Discrimination is almost always the only quantity reported. Calibration is neglected across prediction modelling generally [29], decision-analytic measures such as net benefit are rarer still [30], and TRIPOD+AI asks that discrimination, calibration and intended use be reported together [31]. A feature block can shift predicted risks usefully or improve the ordering of patients within a subgroup without moving a cohort-level AUROC. A study measuring only AUROC cannot separate no added information from added information the metric cannot register. Together, these leave the field with almost no bounded negative results. Nulls are usually unpublished, or stated without an interval, so there is no published estimate of how large a plausible topological effect might be. A well-powered null with an interval attached is therefore more informative than a further positive point estimate.

## 2 Methods

### 2.1 Data source and cohort construction

We obtained data from the National Institute of Allergy and Infectious Diseases (NIAID) TB Portals programme, a multi-country DR-TB surveillance resource. Seven tables were joined on patient and condition identifiers, giving 26,380 longitudinal records across 185 columns for 9,210 patients. Patients contributed a mean of 2.86 records each. Repeated records for one patient are near-duplicate points. If left in the dataset, they inflate local density, shorten nearest-neighbour distances and add spurious zero-persistence structure, distorting the Gower geometry, the Mapper cover and the persistence diagrams simultaneously and unevenly, because follow-up intensity varies with site and severity. Our aggregation removed this artefact before any geometry was computed. We collapsed records to one row per patient by worst-case aggregation. We aggregated binary clinical and imaging indicators by maximum, so a finding was set positive if it was ever recorded. We aggregated numeric burden and severity scores by maximum. We mapped eight ordinal severity variables to numeric ranks and aggregated by maximum rank. We aggregated drug resistance by clinical priority (Sensitive *<* MDR non-XDR *<* Pre-XDR *<* XDR) and outcome by severity priority (Cured *<* Completed *<* Still on treatment *<* Failure *<* Lost to follow-up *<* Palliative care *<* Died). This gave 9,210 patients and 179 features.

We also engineered composite clinical, resistance and radiological features on the training partition only. A cross-validated score combining mutual information, random forest impurity importance and an ANOVA *F* statistic reduced 217 variables (original plus engineered features) to a fixed schema of *d*= 99 clinical features, the baseline set throughout this study.

### 2.2 Outcome definition and analytic cohort

We mapped the eight raw outcomes to three groups. *Positive* is Cured and Completed. *Negative* is Died, Failure, Lost to follow-up and Palliative care. *Pending* is Still on treatment and Unknown. The classification target is binary: Positive against Negative. We excluded Pending patients from classification but deliberately retained them in the Gower dissimilarity, the UMAP embedding and the Mapper graph. An unlabelled patient still occupies a position in clinical feature space. Removing them would thin the manifold non-randomly, since attrition tracks severity, and the recovered geometry would then describe a biased subsample. Pending patients contribute coordinates only. Every label-dependent quantity, including node purity and the derived risk and success scores (defined in Section 2.6.2), is computed over Positive and Negative members alone. A single stratified 80/20 split at patient level gave 7,368 training and 1,842 lockbox patients, with zero overlap enforced by assertion. Of these, 264 and 66 respectively were Pending, leaving 7,104 evaluable training and 1,776 evaluable lockbox patients. The evaluable training cohort is 74.0% Positive and 26.0% Negative. The evaluable lockbox holds 1,313 Positive (73.9%) and 463 Negative (26.1%).

### 2.3 Leakage-controlled pipeline order

We used 10-fold stratified cross-validation, with folds stratified on the ternary treatment-outcome label. The same processing sequence was applied within every cross-validation fold and identically at the training-to-lockbox transition. The sequence is as follows. We first split the data into training and test partitions. We then fit a median imputer on the training patients and apply it to fill missing values in both partitions. Next, we compute pairwise Gower distances over all training patients to form a square distance matrix, and project test patients into this space using only the feature ranges from the training fold. We fit a UMAP embedding on the training Gower matrix and use it as the Mapper lens. We build the Mapper graph on this training embedding, and derive the D^+^ topological features from training-fold outcome labels only, so node purity estimates carry no test information. Simplicial SMOTE was then applied to the training fold to address class imbalance, after which a `StandardScaler` was fitted on the real, non-synthetic training patients and used to scale both matrices. We then fit the classifier on the scaled, balanced training data. We evaluate on the original test fold, which is not resampled and retains the natural class distribution. Fig. 1 marks the training-to-test boundary and indicates which steps involve fitted transformations.

**Fig. 1.**
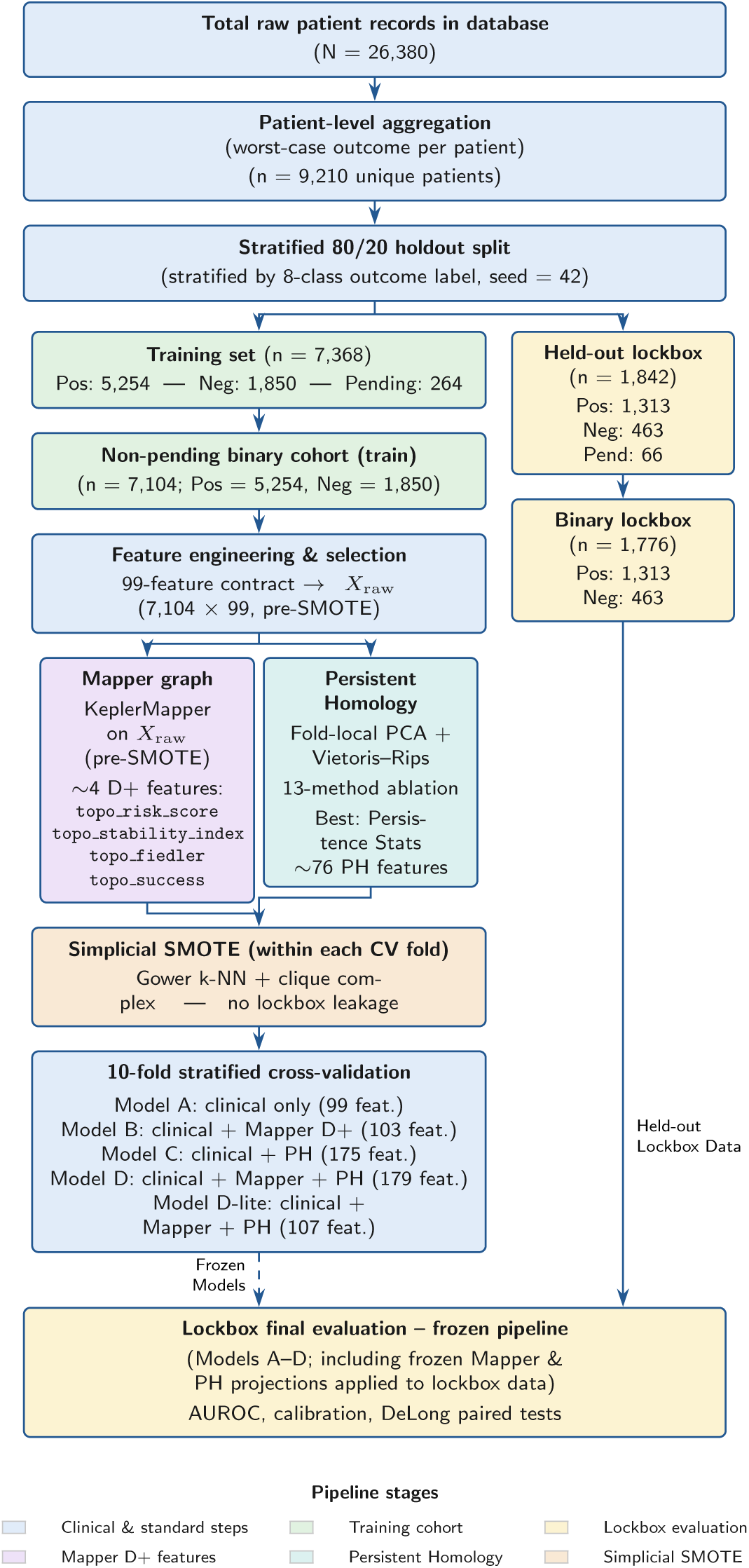
Cohort derivation, feature engineering and evaluation pipeline for DR-TB prediction. Data are initially split into an 80% training set and a 20% held-out lockbox. Model training and topology-based feature generation (Mapper and Persistent Homology) are strictly isolated from the lockbox to prevent data leakage.

Two ordering choices define the design. First, balancing follows topological feature extraction. In an earlier pipeline variant, we applied balancing before Mapper construction and found that synthetic points inflated local density, shifted node membership and altered node purity, distorting the topology the features are meant to measure. Synthetic points are interpolations inside the convex hull of real neighbourhoods, so they change the geometry on which the graph is built. In the current design the graph is built on real training patients only and synthetic patients inherit the D^+^ values of their nearest real training neighbour in the 99-dimensional imputed clinical space by Euclidean 1-NN. Second, the lockbox was written to disk at the split and was not opened, inspected or used in any engineering, selection, tuning or diagnostic step until the single final evaluation.

Our balancing approach used mixed-type Simplicial SMOTE [32], which samples uniformly inside simplices of the clique complex of a symmetric *k*-NN graph rather than along edges. That graph is built on Gower distance, so binary and categorical variables are matched exactly rather than treated as continuous.

### 2.4 Dissimilarity

The feature matrix mixes continuous scores, binary radiological flags and ordinal severity ranks. Euclidean distance treats all of these as continuous and is scale sensitive, so we used the Gower coefficient [25]:

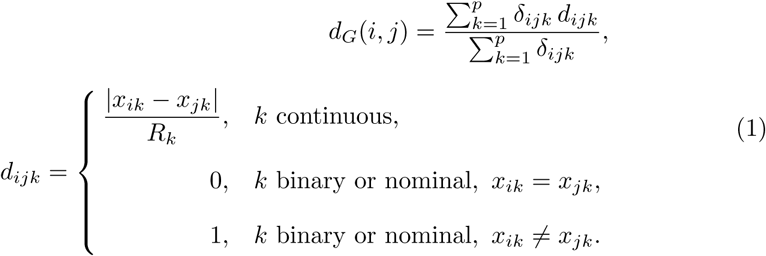

where *δ_ijk_*= 1 if both *x_ik_* and *x_jk_* are observed (and *R_k_ >* 0) and *δ_ijk_*= 0 otherwise, and *R_k_* is the range of feature *k* in the training fold [25]. Continuous features thus contribute a range-normalised absolute difference, binary and nominal features contribute 0 on agreement and 1 on disagreement, and pairs with a missing value are excluded from both numerator and denominator, so the average is taken over jointly observed variables only. The resulting matrix is symmetric with *d_G_*(*i, i*) = 0 and, for values within the training range, lies in [0, 1].

Gower distance serves two roles. First, it defines the *k*-NN graph for Simplicial SMOTE. Second, it is the input to UMAP and hence to the Mapper pipeline. Persistent homology was *not* computed on the Gower dissimilarity but on Euclidean distances in a scaled principal-component space. This distinction matters. The persistence diagrams describe the geometry of that Euclidean embedding, not the mixed-type dissimilarity space, and the two agree only insofar as the embedding preserves Gower structure, which is not guaranteed for binary and ordinal variables. Gower-based persistence, using the dissimilarity matrix as a precomputed filtration, is a stated limitation and a direct extension.

### 2.5 Persistent homology

Persistent homology tracks how topological features of a point cloud appear and disappear as a scale parameter grows [33]. In a Vietoris–Rips filtration, two points are joined when their distance is at most *ɛ*, and a simplex enters when all its vertices are pairwise joined. As *ɛ* grows, components merge and cycles form and fill. Each feature is recorded as a birth-death pair, and the collection of pairs in dimension *q* is the degree-*q* persistence diagram. *H*_0_ records connected components, *H*_1_ records one-dimensional cycles.

We designed the analysis to be local and per-patient. We standardised features with a `RobustScaler` and reduced dimensionality with PCA, keeping the smallest number of components reaching 85% cumulative explained variance, capped at 25. For each patient, we formed a local cloud from that patient plus its 25 Euclidean nearest neighbours in the training PCA space, giving 26 points. For lockbox patients, the 25 neighbours were drawn exclusively from the training set. We computed Rips persistence on each cloud with `ripser` [34, 35] at maxdim = 1, yielding *H*_0_ and *H*_1_ only.

We opted for the local construction approach for this problem instead of a global one. Running a single Rips filtration over all 7,104 non-pending training patients is not feasible: the distance matrix grows as *O*(*n*^2^) in memory, and the simplex count grows combinatorially with patient count. Replacing this with one small filtration per patient, each over 26 points, makes the computation tractable. This design changes what the descriptor measures. A local filtration produces a patient-level quantity: it describes the topological shape of an individual patient’s clinical neighbourhood. A cohort-level persistence diagram describes the whole population simultaneously and cannot be assigned to an individual patient.

*H*_0_ bars of zero persistence (coincident points) were excluded: Ripser omits them, and in addition all diagrams were filtered to finite bars with death *>* birth. For a 26-point local Vietoris–Rips cloud the maximum number of finite *H*_0_ bars is 25; the observed mean of 24.9 (range 22–25) lies at this ceiling because the count is fixed by construction and falls below 25 only when patients share identical PCA coordinates. The *H*_0_ bar count therefore carries almost no discriminative information, and *H*_0_ information in the model is carried by the lifespan-based features (mean, variance, entropy).

Three limitations follow from this design. First, we computed homology only up to dimension one, capturing connected components (*H*_0_) and loops (*H*_1_); two-dimensional voids (*H*_2_) were not computed and are not represented in any model feature. Second, we used no global filtration over the full cohort for modelling. Third, distance to measure (DTM) weighted filtrations and Gower-based persistence were not evaluated, as neither would have materially altered the evaluation design or the primary conclusion.

### 2.6 Mapper and the D^+^ features

Mapper [18] summarises a point cloud as a graph. A lens maps the data to a low-dimensional space, that space is covered by overlapping sets, points in each pre-image are clustered, each cluster becomes a node and nodes sharing a point are joined. Overlap is what preserves connectivity a hard partition would destroy. The lens is the first two dimensions of a UMAP embedding [27] with *n* neighbors = 15 and min dist = 0.1, fitted on the training-fold Gower matrix under a fixed seed. Unseen patients are projected by distance-weighted interpolation of the UMAP coordinates of their *k* = 5 nearest training patients under Gower distance; this approximation is used in place of the standard UMAP transform because Mapper requires 2-D coordinates rather than a parametric embedding. The cover uses 10 hypercubes per axis at 50% overlap. Pre-images are clustered with DBSCAN [36] at *ε* = 0.3 and min samples = 3. We built graphs with KeplerMapper [37].

#### 2.6.1 Hyperparameter stability ablation

Mapper parameters were fixed by a stability criterion rather than by a downstream classification metric. This choice is necessary because the Mapper graph feeds directly into the classifier. Selecting parameters by AUROC or F1 would allow the downstream performance measure to influence the input features, which is circular.

We evaluated a 3 × 3 grid of nine configurations: *n*_cubes_ ∈ {8, 10, 12} crossed with *ε* ∈ {0.2, 0.3, 0.4}, with overlap fixed at *ρ* = 0.5 and min_samples = 3. For each configuration, *B* = 20 stratified bootstrap resamples were drawn from 80% of the fold-1 training UMAP embedding. On each resample, a Mapper graph was constructed and two stability metrics were recorded:

1. **Node-count coefficient of variation** CV*_n_* = 100 × *σ_nB_ /µ_nB_*, where *µ_nB_* and *σ_nB_* are the mean and standard deviation of node counts across the *B* resamples.
2. **Purity signal standard deviation** 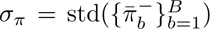, where 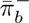 is the mean negative purity across all nodes of the *b*-th bootstrap graph.

Both thresholds, CV*_n_ <* 15% and *σ_π_ <* 0.05, were fixed before the ablation was run. All nine configurations satisfied both criteria. CV*_n_* ranged from 3.8% to 7.9% and *σ_π_* ranged from 0.010 to 0.023 across the grid. The selected configuration (*n*_cubes_ = 10, *ε* = 0.3, *ρ* = 0.5) yielded CV*_n_*= 6.5% and *σ_π_*= 0.023.

Because all nine configurations passed, the ablation does not identify a single optimal setting. The topological structure recovered by Mapper is stable across the full parameter range and is not an artefact of any particular configuration.

#### 2.6.2 Variant D^+^ topological feature extraction

For patient *i*, let N (*i*) denote the set of Mapper nodes to which that patient belongs, where *n_i_* = |N (*i*)| is the number of such nodes. A patient may belong to more than one node because the cover elements overlap. For each node *v* ∈ N (*i*), *π^−^*(*v*) denotes negative purity, *d*_fail_(*v*) the hop distance to the nearest high-risk node, *d*_success_(*v*) the hop distance to the nearest high-success node, and *m_v_* = |M(*v*)| the node size, where M(*v*) is the set of patients assigned to node *v*. Four patient-level features were derived from the Mapper graph and together form the Variant D^+^ block:

i. *topo_risk_score:* the mean of *π^−^*(*v*)*/*(1 + *d*_fail_(*v*)) over N (*i*).
ii. *topo_success_score:* the mean of 1*/*(1 + *d*_success_(*v*)) over N (*i*).
iii. *topo_stability_index:* the mean node size, discounted by multiplicity and branch proximity:

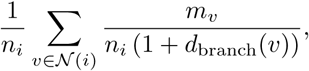 where *d*_branch_(*v*) is the hop distance from *v* to the nearest branch node (degree ≥ 3), set to 0 when the graph contains no branch node. The factor 1*/n_i_* in the denominator is a multiplicity discount: patients assigned to many overlapping nodes receive proportionally lower values, down-weighting patients assigned to densely overlapping cover elements.
iv. *topo_fiedler:* the mean of the Fiedler value *ψ*(*v*) over N (*i*), where *ψ*(*v*) is the entry of the Fiedler vector at node *v*, defined below.

All purities and thresholds were computed from training labels only. Node purity values used to construct each training patient’s D^+^ features were computed from the outcomes of all other training patients assigned to that node, excluding the patient itself, so each patient’s own outcome label does not contribute to its own feature values. Persistent homology features for synthetic SMOTE patients were assigned by the same Euclidean 1-NN procedure used for D^+^ values: each synthetic patient inherited the PH features of its nearest real training neighbour in the 99-dimensional imputed clinical space. The *topo_stability_index* is min–max normalised to [0, 1] using the minimum and maximum observed in the training set, which are frozen before application to test patients. Test values are *not* clipped: a test patient whose raw value falls outside the training range receives a normalised value outside [0, 1]. We computed *topo_fiedler* as follows. Let **A** be the binary adjacency matrix of the training-fold Mapper graph, with *A_uv_* = 1 if nodes *u* and *v* share at least one patient and *A_uv_* = 0 otherwise, and let **D** be the diagonal matrix of node degrees, *D_vv_* = Σ*_u_ A_vu_*. The graph Laplacian is **L** = **D**−**A**. Its second-smallest eigenvalue *λ*_2_ is the algebraic connectivity of the graph [38]: *λ*_2_ = 0 if and only if the graph is disconnected, and the associated eigenvector is the Fiedler vector. Because **L1** = **0**, the all-ones vector **1** is always an eigenvector for eigenvalue zero, so the Fiedler vector is orthogonal to **1** and therefore sums to zero over the graph.

When the Mapper graph has *k* ≥ 2 connected components, **L** is block-diagonal: after permuting rows and columns by component membership, it takes the form diag(**L**_1_, **L**_2_*, …,* **L***_k_*), where **L***_c_* is the graph Laplacian of the subgraph induced by component *C_c_*. Consequently *λ*_2_(**L**) = 0 and a global Fiedler vector is not uniquely defined. We therefore compute the Fiedler vector separately within each component. For each component *C_c_* with |*C_c_*| ≥ 2, let *ψ_c_* be the unit-norm eigenvector of **L***_c_* corresponding to its own algebraic connectivity *λ*_2_(**L***_c_*) *>* 0 [38]; each node *v* ∈ *C_c_* takes *ψ*(*v*) = *ψ_c_*(*v*). Nodes in singleton components (|*C_c_*| = 1) take *ψ*(*v*) = 0. The computation uses the training-fold graph only and no outcome labels. A patient’s *topo fiedler* value is the unweighted mean of *ψ*(*v*) over the nodes they are assigned to.

For training patients, all assigned nodes lie within one component, because Mapper places an edge between any two nodes that share a patient, so patients sharing a node are always in the same component. A held-out patient is assigned to the single nearest node by centroid distance in the UMAP projection; if the implementation assigns more than one node, the mean is taken over the corresponding component-local entries. Because each *ψ_c_* has unit norm on *C_c_* and sums to zero on *C_c_*, entries are comparable within a component but not across components: a component with many nodes has smaller individual entries by normalisation than a component with few nodes. For patients whose nodes span different connected components, *topo_fiedler* summarises local spectral position within each component rather than a coordinate in a single global spectral embedding. It is therefore used here as a patient-level descriptor for prediction, and values should not be compared as positions in a shared embedding space.

### 2.7 Vectorisation ablation

A persistence diagram is a multiset, not a vector, so it must be mapped to fixed-length coordinates before a classifier can use it. The map is not neutral and the literature offers no default, so we treated the choice as a designed ablation over the taxonomy of [23] rather than a search for a winning number. We implemented all 13 catalogued methods: persistence statistics, entropy summary, algebraic functions, tropical coordinates, complex polynomial, Betti curve, lifespan curve, persistence landscape, persistence silhouette, persistence image, template functions, adaptive templates and ATOL. These fall into statistical, algebraic, curve, functional and ensemble families, which between them span the ways a diagram can be summarised: by moments, by algebraic invariants, by functions of the filtration parameter, by kernel density on the birth-death plane, and by learned landmarks.

We applied each method separately to *H*_0_ and *H*_1_ and concatenated the outputs. Three points govern leakage. The data-driven methods, adaptive templates and ATOL, learn template centres, covariances and landmarks, and were refitted inside each training fold. Curve-type methods need a fixed evaluation grid, so *t*-ranges were fixed from training barcodes and applied unchanged to test barcodes. The ablation used 5-fold cross-validation across the four-classifier panel. Persistence statistics performed best and we carried it forward to all downstream analysis.

### 2.8 Model configurations and classifiers

Five configurations were compared, all sharing the 99-feature clinical baseline (Table 1). Model D-lite compresses the persistence block: within a fit-only subset of the training data, PH columns were filtered by variance, ranked by point-biserial correlation with the binary outcome, pruned greedily at |*r*| ≥ 0.90, and the retained count *k*_PH_ chosen by inner 3-fold cross-validation over {4, 8, 12, 16}.

**Table 1.** Model configurations compared in this study.

| Model | Feature set | $d$ |
| --- | --- | --- |
| A | Clinical only (baseline) | 99 |
| B | A + Mapper D<sup>+</sup> | 103 |
| C | A + persistence-statistics PH | 175 |
| D | B + persistence-statistics PH | 179 |
| D-lite | B + compressed PH block ($k_{\text{PH}} = 4$) | 107 |

All models share the same 99 clinical features and the same classifier registry. $d$ , number of features.

We used a fixed registry of four classifiers at every stage: logistic regression, random forest [39], support vector machine with an RBF kernel [40] and XGBoost [41]. We applied different cross-validation depth per experiment by design. The A against B Mapper comparison used 10-fold stratified cross-validation, chosen for paired-test power, since the effect under test was expected to be small and paired fold-level contrasts need enough folds to be informative. The PH model benchmark used 5 folds, because each fold recomputes every patient barcode. The 13-method ablation also used 5 folds across the four-classifier panel.

### 2.9 Evaluation protocol

In the primary lockbox analysis, we performed final binary evaluation on the 1,776 non-pending lockbox patients. We partitioned the non-pending training data into three disjoint subsets: a fit set (80%), a calibration set (10%), and a threshold-tuning set (10%). We trained base classifiers on the fit subset, then calibrated probabilities on the calibration subset using Platt scaling [42]:

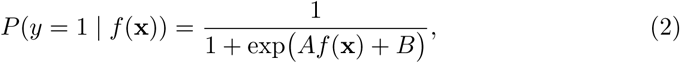

where *A* and *B* are fitted by maximum likelihood on the calibration subset, with *A* typically negative for margin-based classifiers. We selected the operating threshold on the threshold-tuning subset by maximising the Youden index [43]. No subset was used for more than one purpose.

We compared discrimination with paired DeLong tests [28] on the calibrated lockbox probabilities, per classifier. We declared two correction families in advance. The *A-referenced family* holds four comparisons (A against B, C, D and D-lite), Bonferroni corrected at *α*_corr_ = 0.05*/*4 = 0.0125. Bonferroni correction was applied within each classifier’s four comparisons rather than globally across all sixteen, on the grounds that each classifier constitutes an independent evaluation of the same topological augmentation question. A global correction across sixteen tests would have been *α*_corr_ = 0.05/16 = 0.003125, which is more conservative than the design warrants. Equivalently, adjusted *p*-values are *p*_adj_ = min(1, 4*p*) and a comparison is declared significant when *p*_adj_ *<* 0.05.

Bonferroni correction was used rather than false discovery rate (FDR) control. FDR is suited to screening studies where a small fraction of false positives is acceptable [44]. Here, four prespecified comparisons each ask whether a named augmentation improves on the clinical baseline. A single false positive would incorrectly support the central claim of the paper. Bonferroni controls the probability that any one comparison is wrongly declared significant. It is conservative, and that conservatism is appropriate when one false positive is one too many. As a sensitivity check, applying a global Bonferroni correction across all sixteen tests would give *α*_corr_ = 0.05*/*16 = 0.003125. Under that stricter threshold, the three logistic regression comparisons significant at the per-classifier level (against Models B, D and D-lite) remain significant, as their raw *p*-values are below 0.003125; the XGBoost comparison against Model D (*p*_adj_ = 0.0211 under per-classifier correction) does not.

Discrimination alone is insufficient under 74/26 imbalance. AUROC is invariant to prevalence and can stay flat while the ranking of the adverse minority shifts, and it says nothing about whether predicted probabilities are usable as risk estimates [45]. We therefore report AUROC with 95% confidence intervals alongside adverse-class PR-AUC, the Brier score against the base-rate Brier of 0.1927, and sensitivity and specificity at the Youden threshold.

## 3 Results

### 3.1 Cohort characteristics

Table 2 shows the composition of each partition. Lockbox discrimination and calibration analyses used a binary regrouping of the outcome. Of the 1,842 lockbox patients, 1,776 were eligible. The 66 patients recorded as still on treatment or unknown were excluded.

**Table 2.** Cohort composition by partition.

| Class / group | Training | Lockbox | All |
| --- | --- | --- | --- |
| Total patients | 7368 | 1842 | 9210 |
| *Ternary outcome, n (%)* |  |  |  |
| Positive (cured/completed) | 5254 (71.3) | 1313 (71.3) | 6567 (71.3) |
| Negative (died/failure/Lost to follow up/Palliative care) | 1850 (25.1) | 463 (25.1) | 2313 (25.1) |
| Pending (still on treatment/unknown) | 264 (3.6) | 66 (3.6) | 330 (3.6) |
| *Lockbox binary analysis, n (%)* |  |  |  |
| Positive | — | 1313 (73.9) | — |
| Negative | — | 463 (26.1) | — |
| Excluded | — | 66 (3.6) | — |
| Total eligible | — | 1776 | — |

Percentages are computed within each column. The ternary outcome mapping is defined in Section 2.

### 3.2 Vectorisation ablation

We computed local Vietoris–Rips filtrations on a seven-component PCA embedding, which accounted for 85.4% of cumulative explained variance. The resulting *H*_0_ diagrams held a mean of 24.9 finite bars per patient, with a range of 22 to 25. The *H*_1_ diagrams held a mean of 4.6 bars per patient, with a range of 0 to 16. About 2% of patients yielded no *H*_1_ bars.

We computed all thirteen vectorisation methods catalogued by [23] from these diagrams and benchmarked them under five-fold cross-validation using topological features alone, with no clinical features present. Table 3 reports the ranking. Best AUROC ranged from 0.5394 for Persistence Landscape to 0.5829 for Persistence Statistics. Persistence Statistics ranked first, ATOL second at 0.5788, and Adaptive Templates third at 0.5779. Curve-family methods occupied four of the bottom five ranks (9, 11, 12, and 13). XGBoost supplied the best AUROC for 8 of the 13 methods, SVM for 3, and Random Forest for 2. Persistence Statistics was used as the PH representation in all subsequent models.

**Table 3.** Persistence-diagram vectorisation methods ranked by out-of-fold macro-AUROC.

| Rank | Method | Dim. | Family | AUROC | Clf. |
| --- | --- | --- | --- | --- | --- |
| 1 | Persistence Statistics | 76 | Statistical | 0.5829 | RF |
| 2 | ATOL | 20 | Ensemble | 0.5788 | XGBoost |
| 3 | Adaptive Templates | 20 | Ensemble | 0.5779 | XGBoost |
| 4 | Tropical Coordinates | 14 | Algebraic | 0.5710 | XGBoost |
| 5 | Template Functions | 98 | Functional | 0.5709 | RF |
| 6 | Algebraic Functions | 8 | Algebraic | 0.5664 | XGBoost |
| 7 | Entropy Summary | 100 | Statistical | 0.5644 | SVM |
| 8 | Complex Polynomial | 20 | Algebraic | 0.5635 | XGBoost |
| 9 | Lifespan Curve | 100 | Curve | 0.5622 | XGBoost |
| 10 | Persistence Image | 200 | Functional | 0.5516 | SVM |
| 11 | Persistence Silhouette | 100 | Curve | 0.5435 | XGBoost |
| 12 | Betti Curve | 100 | Curve | 0.5415 | SVM |
| 13 | Persistence Landscape | 500 | Curve | 0.5394 | XGBoost |

Methods are ranked by best out-of-fold one-versus-one macro-AUROC across the classifier panel. Features are topological only, with no clinical covariates. Five-fold cross-validation. Dim., dimensionality of the vectorised representation (7104 samples × $d$ features); Clf., best classifier; RF, random forest; SVM, support vector machine.

### 3.3 Mapper feature redundancy assessment

The four Mapper D^+^ features showed low mutual redundancy. The largest variance inflation factor was 3.318, for the topological stability index, against a flagging threshold of 5.0. The largest absolute pairwise Pearson correlation was 0.600, between the topological risk score and the topological success score. No pair exceeded the |*r*| *>* 0.60 4 flagging threshold.

### 3.4 Primary comparison on the held-out lockbox

No topologically augmented configuration achieved a meaningful improvement in lockbox AUROC over the clinical-only baseline under any of the four classifiers. One 2 comparison (SVM, Model B) met the corrected significance threshold in the augmented model’s favour, but by only +0.0005 AUROC units. Table 4 reports calibrated discrimination for the five configurations across the four classifiers, together with the DeLong paired comparison against Model A. The highest AUROC among the 20 model and classifier combinations was 0.7548, achieved by Model A with XGBoost. Model A with XGBoost also gave both the best adverse-class PR-AUC at 0.5605 and the lowest Brier score at 0.1580, against a base-rate Brier of 0.1927. The highest-AUROC augmented configuration was Model C with Logistic Regression at an AUROC of 0.7519.

**Table 4.** Calibrated lockbox AUROC and DeLong paired comparisons against Model A.

| Model | Clf. | AUROC | $\Delta$ [95% CI] | $p_{\text{adj}}$ |
| --- | --- | --- | --- | --- |
| A (99) | LR | 0.7523 | reference | — |
|  | RF | 0.7345 | reference | — |
|  | SVM | 0.6906 | reference | — |
|  | XGB | 0.7548 | reference | — |
| B (103) | LR | 0.7351 | −0.0172 [−0.025, −0.010] | < 0.0001* |
|  | RF | 0.7401 | +0.0056 [−0.003, +0.014] | 0.8040 |
|  | SVM | 0.6911 | +0.0005 [+0.000, +0.001] | 0.0478* |
|  | XGB | 0.7475 | −0.0073 [−0.014, −0.001] | 0.1184 |
| C (175) | LR | 0.7519 | −0.0004 [−0.005, +0.004] | 1.0000 |
|  | RF | 0.7188 | −0.0156 [−0.033, +0.001] | 0.2859 |
|  | SVM | 0.6906 | −0.0000 [−0.004, +0.004] | 1.0000 |
|  | XGB | 0.7439 | −0.0109 [−0.020, −0.002] | 0.0889 |
| D (179) | LR | 0.7370 | −0.0153 [−0.023, −0.008] | 0.0003* |
|  | RF | 0.7162 | −0.0182 [−0.035, −0.001] | 0.1351 |
|  | SVM | 0.6910 | +0.0004 [−0.004, +0.004] | 1.0000 |
|  | XGB | 0.7411 | −0.0137 [−0.023, −0.004] | 0.0211* |
| D-lite (107) | LR | 0.7312 | −0.0210 [−0.029, −0.013] | < 0.0001* |
|  | RF | 0.7322 | −0.0023 [−0.013, +0.008] | 1.0000 |
|  | SVM | 0.6911 | +0.0005 [−0.000, +0.001] | 0.3746 |
|  | XGB | 0.7467 | −0.0081 [−0.016, −0.001] | 0.1468 |

Calibrated lockbox AUROC for five model configurations across four classifiers, with DeLong paired comparisons against Model A. Bonferroni correction over four A-referenced tests per classifier: $p_{\text{adj}} = \min(1, 4p)$ , significant when $p_{\text{adj}} < 0.05$ (equivalently $p < \alpha_{\text{corr}} = 0.05/4 = 0.0125$ ). $n = 1776$ ( $n_{\text{pos}} = 1313$ , $n_{\text{neg}} = 463$ ). Base-rate Brier score = 0.1927. Model A is the reference; $\Delta$ and CI are not applicable. Feature counts in parentheses. A, clinical only; B, clinical + Mapper D<sup>+</sup>; C, clinical + PH; D, clinical + PH + Mapper D<sup>+</sup>; D-lite, clinical + Mapper D<sup>+</sup> + 4 compressed PH dimensions. LR, logistic regression; RF, random forest; SVM, support vector machine; XGB, XGBoost. $\Delta$ is the augmented model’s AUROC minus Model A’s AUROC for the same classifier.
\* Significant after Bonferroni correction ( $p_{\text{adj}} < 0.05$ ). Four of the five favour Model A; the SVM comparison with Model B favours the augmented model by +0.0005 AUROC, with an interval of near-zero width (plotted at scale in Fig. 2).

Sixteen A-referenced DeLong tests were run, four per classifier, with Bonferroni correction (adjusted *p*-values are four times the raw *p*-values, capped at 1, and are compared with 0.05; equivalently, *α*_corr_ = 0.0125 for raw values). Four of the sixteen point estimates were positive: Model B with Random Forest (+0.0056, 95% CI −0.003 to +0.014; *p*_adj_ = 0.8040) and Models B, D and D-lite with SVM (+0.0005, +0.0004 and +0.0005 respectively). Five tests met the corrected threshold. Four favoured Model A: Logistic Regression against Model B (Δ = −0.0172, 95% CI −0.025 to −0.010; *p*_adj_ *<* 0.0001), against Model D (Δ = −0.0153, 95% CI −0.023 to −0.008; *p*_adj_ = 0.0003) and against Model D-lite (Δ = −0.0210, 95% CI −0.029 to −0.013; *p*_adj_ *<* 0.0001), and XGBoost against Model D (Δ = −0.0137, 95% CI −0.023 to −0.004; *p*_adj_ = 0.0211). The fifth favoured the augmented model: SVM with Model B (Δ = +0.0005, 95% CI +0.000 to +0.001; *p*_adj_ = 0.0478). Its interval is very narrow because the two SVM models rank patients almost identically, and a difference of this size has no practical meaning.

The largest upper confidence limit among the sixteen tests was +0.014 AUROC units (Random Forest, Model B); every other upper limit was at or below +0.008 (Fig. 2). Interval widths for the twelve non-SVM tests ranged from 0.009 to 0.034 AUROC units. The four SVM intervals were narrower, at 0.001 to 0.008 units.

**Fig. 2.**
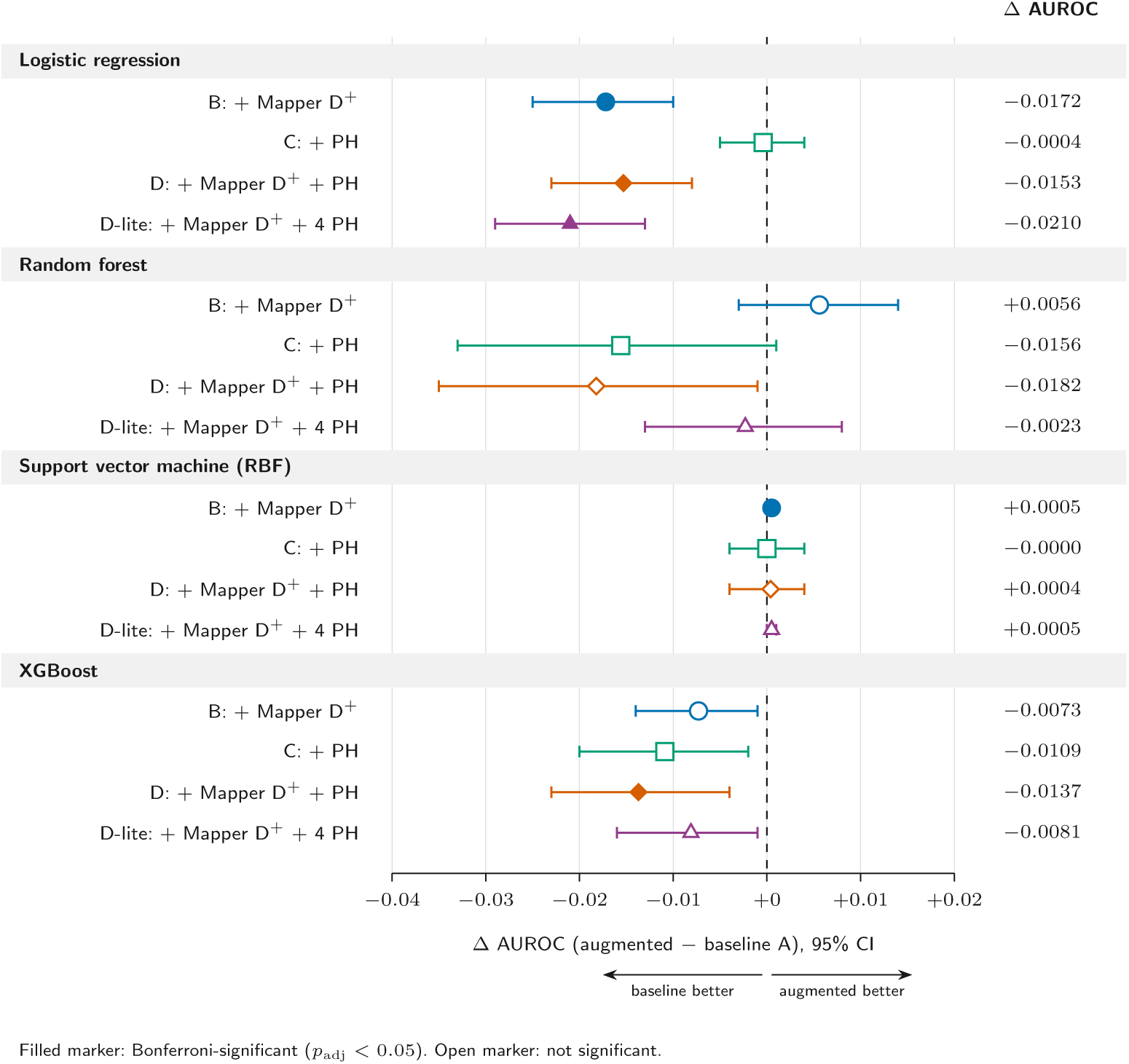
DeLong paired AUROC differences against the clinical-only baseline, by classifier. Each augmented configuration is compared with Model A, with 95% confidence intervals, grouped by classifier. Filled markers are significant after Bonferroni correction over four comparisons per classifier (*p*_adj_ *<* 0.05, equivalently *p < α*_corr_ = 0.0125). Points to the left of the dashed zero line indicate that the augmented model performed worse than the baseline. The largest upper confidence limit is +0.014 (Random Forest, Model B). All four A-referenced comparisons per classifier (Models B, C, D and D-lite) are shown; numerical values for all sixteen tests appear in Table 4

Operating-point behaviour varied by configuration. At the Youden-optimised threshold, sensitivity ranged from 0.4174 for Model C with SVM to 0.7631 for Model B with XGBoost, and specificity ranged from 0.5832 for Model D-lite with SVM to 0.8143 for Model C with SVM.

Table 5 reports the lockbox macro-F1 scores at each model’s tuned decision threshold. Macro-F1 ranged from 0.516 to 0.661 across the 20 model–classifier combinations, and no feature set was consistently best. The highest value was obtained by Model C with Logistic Regression (0.661), closely followed by Model B with XGBoost (0.661) and D-lite with XGBoost (0.658). The clinical baseline (Model A) was strongest with Random Forest (0.651) and did not rank first for any classifier. The effect of topological augmentation depended on the classifier. Adding Mapper D^+^ features (Model B) raised XGBoost macro-F1 from 0.635 to 0.661, while the persistent homology block (Model C) raised Logistic Regression macro-F1 from 0.607 to 0.661. The same blocks lowered other classifiers: the SVM (RBF) macro-F1 fell from 0.614 for Model A to 0.516 for Models C and D. Because macro-F1 depends on the tuned threshold, these differences reflect operating-point trade-offs rather than consistent gains in discrimination.

**Table 5.** Lockbox macro-F1 by model and classifier at the tuned decision threshold (*n* = 1,776 non-pending patients). The highest value in each column is shown in bold.

| Model | Logistic Regression | Random Forest | SVM (RBF) | XGBoost |
| --- | --- | --- | --- | --- |
| A | 0.6073 | **0.6509** | 0.6142 | 0.6346 |
| B | 0.6024 | 0.6390 | 0.6142 | **0.6607** |
| C | **0.6614** | 0.6325 | 0.5163 | 0.5812 |
| D | 0.5929 | 0.6056 | 0.5168 | 0.6329 |
| D-lite | 0.5902 | 0.6430 | **0.6148** | 0.6575 |

### 3.5 SHAP feature attribution

SHAP (SHapley Additive exPlanations) decomposes each model’s lockbox prediction into additive per-feature contributions grounded in cooperative game theory [46]. For patient *i* and model output *f*^^^(*x_i_*), the decomposition satisfies the efficiency axiom:

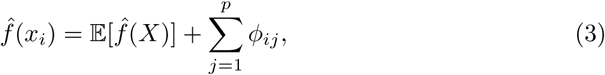

where *ϕ_ij_* is the Shapley value of feature *j* for patient *i*, and E[*f*^^^(*X*)] is the baseline expected model output over the training distribution. This analysis is applied exclusively to the held-out lockbox. Positive SHAP values push predictions toward the positive-outcome class (Cured / Completed); negative values push toward adverse outcomes. SHAP values were computed on the uncalibrated base-classifier output for Logistic Regression, Random Forest, and XGBoost (log-odds scale for Logistic Regression and XGBoost; probability scale for Random Forest). For the SVM, SHAP was applied to the Platt-calibrated probability using KernelExplainer. Platt scaling is a monotone transformation of the model output, so it leaves the ranking of patients unchanged. However, because the sigmoid is nonlinear, it does not exactly preserve the magnitude or ranking of per-feature attributions. Block-level conclusions therefore rely on within-model fractions of total |*ϕ*| rather than on absolute cross-model SHAP magnitudes.

Clinical features accounted for 87% and 89% of total mean absolute SHAP contribution in Models B and C respectively, while Mapper D^+^ contributed 13% in Model B and persistent homology 11% in Model C (Fig. 3b). In Model D, the combined topological contribution was 35% of total attribution (11% Mapper D^+^; 24% PH). In D-lite, clinical features accounted for 80%, with Mapper D^+^ contributing 12% and PH 8%. Per feature, the Mapper D^+^ attributions (0.089 to 0.092) were roughly four times those of the average clinical feature in the same model (0.022 to 0.025; Fig. 3a), so the modest block shares reflect the small size of the block, four features against 99, rather than weak individual attributions. These proportions indicate that TDA features supplemented rather than replaced conventional clinical predictors. Because the D^+^ scores are outcome-informed summaries of the training cohort (Section 2.6.2), high attribution is expected and is not evidence of independent predictive value.

**Fig. 3.**
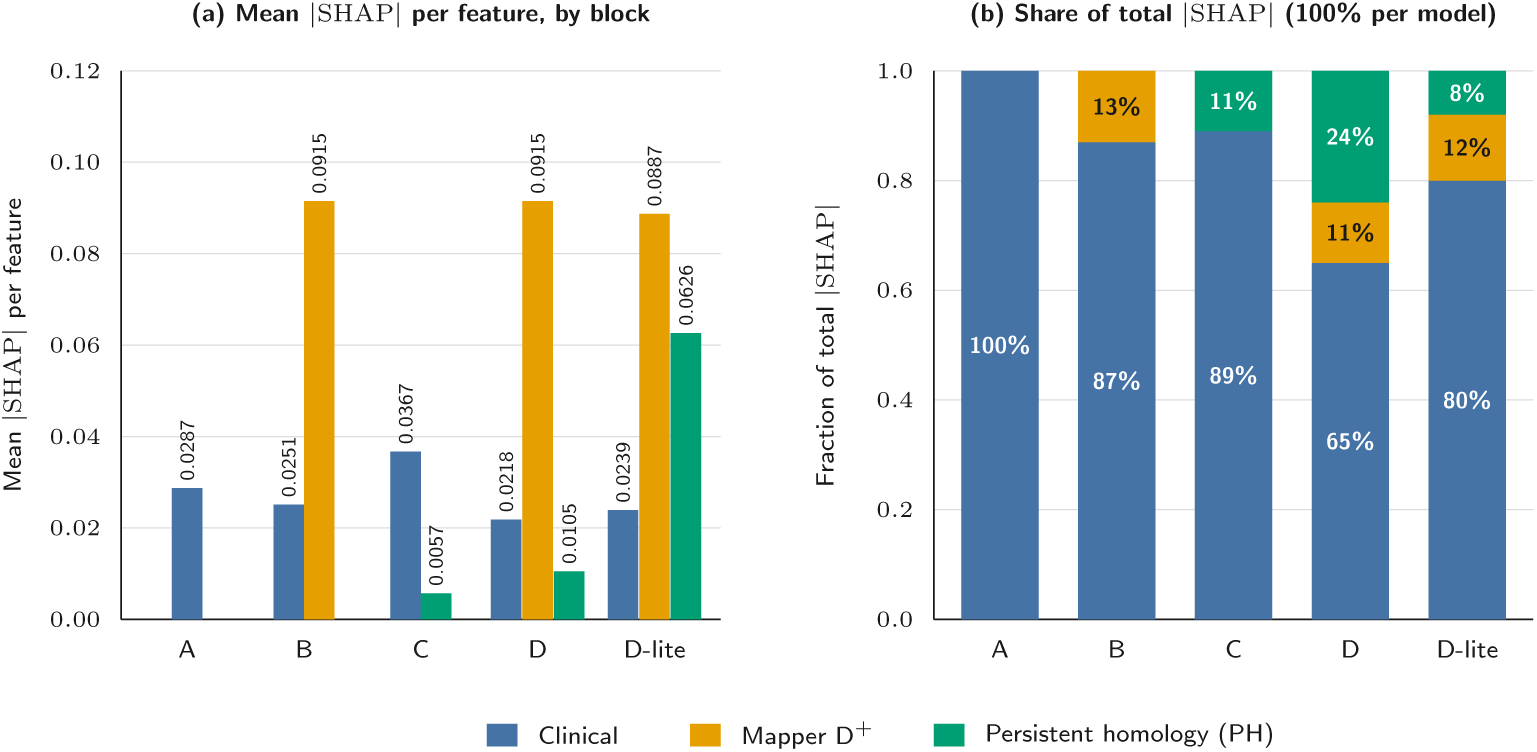
Cross-model SHAP feature-block attributions on the held-out lockbox set. (a) Mean absolute SHAP value per feature within each block (clinical, 99 features; Mapper D^+^, 4; persistent homology, 76 in Models C and D and 4 in D-lite), with values printed above the bars. (b) Each block’s share of total mean absolute SHAP attribution, summing to 100% per model. Clinical variables remained the dominant source of attribution in every model, but TDA-derived features provided a complementary signal, with the largest combined non-clinical contribution (35%) in Model D.

## 4 Discussion

We asked what topological structure adds to a clinical prediction model and how that addition should be measured. The discriminative answer is negative, and the structural one is not.

### 4.1 Interpreting the null result

Our AUROC values ranged from 0.69 to 0.75, in line with the 0.71 to 0.74 reported for models using baseline clinical data [2, 47]. Higher pooled estimates exist, but they come predominantly from studies with limited external validation and a high risk of bias [48]. DR-TB outcomes are shaped by comorbidities such as HIV and alcohol misuse [49] and by treatment adherence [50]. Three limits bound what the design can establish. The design is powered for moderate AUROC differences on 1,776 lockbox patients, of whom 463 have adverse outcomes. Differences of a few thousandths of AUROC lie within the noise of a test set this size, so the study can rule out a useful discrimination gain but cannot resolve a marginal one. The analysis is single-cohort. Splitting one TB Portals extract gives an internal held-out estimate, not external validity. Site composition, imaging platforms and outcome ascertainment are shared across the split, so transportability to another programme is untested. A null DeLong result does not establish equivalence. Failing to reject equal AUROC is not evidence that AUROCs are equal. Equivalence requires a prespecified margin and an equivalence test, neither of which we undertook. Where we report that topological augmentation gives no discrimination gain, the claim is that none was detected under a protocol built to detect one, not that none exists.

No topologically augmented configuration meaningfully improved lockbox AUROC over the clinical-only baseline for any of the four classifiers (Section 3.4). Across all sixteen DeLong comparisons the upper limit of the 95% confidence interval never exceeded +0.014 AUROC (Random Forest, Model B), and every other upper limit was at or below +0.008. That is a real constraint: an improvement of the size usually claimed for a new feature block in clinical prediction would have been detected. Under the support vector machine the constraint is tighter still, with all four intervals at or below 0.008 AUROC in width and all four point estimates within 0.0005 of zero. One of these, SVM with Model B, was nominally significant at the adjusted p-value level (*p*_adj_ = 0.0478) but without any practical meaning.

The constraint is asymmetric. The widest intervals, random forest against Model C and against Model D, each spanned 0.034 AUROC units. For the Model C comparison the data exclude improvement beyond +0.001 but remain compatible with a decrement of moderate size. The study therefore supports a bounded negative claim, not a symmetric one: for this cohort, this baseline and these classifiers, topological augmentation does not improve rank discrimination by more than 0.014 AUROC units (the largest upper limit of all sixteen 95% confidence intervals), and in several configurations it reduces it. Four comparisons, three under logistic regression and one under XGBoost, showed a significant reduction.

Two qualifications limit this conclusion. The study was not designed as a formal equivalence trial; no margin was prespecified and the lockbox size was fixed by the available data. Second, the null is conditional on this baseline: the 99-feature clinical schema reached an AUROC of 0.7548 (XGBoost; 0.7523 with logistic regression) and the best adverse-class PR-AUC and Brier score of any configuration tested, and a well-specified baseline is the hardest condition under which to show incremental value.

### 4.2 Discriminative metrics as the wrong instrument

AUROC is a rank statistic computed over the entire cohort. Two structural properties matter for evaluating any new feature block. First, AUROC is dominated by the majority of patient pairs. A representation that sharpens risk estimates for a small subgroup leaves most pairwise rankings unchanged, and its contribution to the cohort-wide statistic is diluted proportionally. If topological position carries information mainly about patients in unusual regions of the data manifold, those patients are by definition few, and their effect on AUROC will be correspondingly small. Second, AUROC is insensitive to where on the risk scale a change occurs. A model that reorders patients near a decision threshold is more useful than one that reorders patients who are already clearly low risk, yet AUROC assigns equal weight to both [30].

A pattern in the results is consistent with this argument. Model C with Logistic Regression achieves the highest macro-F1 of all 20 model-classifier combinations (0.6614) and a sensitivity of 0.7113, against 0.5636 for the same classifier on the clinical baseline, while its AUROC (0.7519) is statistically indistinguishable from the baseline (0.7523). The Youden threshold was fixed on the held-out tuning partition, not on the lockbox, so this is not threshold selection on test data. Two interpretations are consistent with the evidence. The sensitivity shift may reflect genuine enrichment of the classifier’s probability estimates near the decision boundary by the persistent homology feature block. Equally, the same pattern arises if the Youden threshold simply lands at a different operating point on an otherwise unchanged ROC curve. The data collected here cannot separate these interpretations, because neither calibration within risk strata nor decision-analytic comparisons were prespecified as primary endpoints.

Separating the two interpretations requires measurements designed for that purpose. Calibration should be assessed within risk strata rather than only through an aggregate Brier score. Discrimination should be examined conditional on baseline model uncertainty. Comparisons should be decision-analytic, reporting net benefit across a range of plausible threshold probabilities [30].

Work that claims structural value for a topological representation should prespecify measurements capable of detecting it. Testing only a global rank statistic when the hypothesised benefit is local and subgroup-specific is a mismatch between the instrument and the claim. Future evaluations of TDA-augmented clinical prediction models should include calibration-stratified and decision-analytic endpoints as primary outcomes alongside AUROC, specified before outcome data are accessed.

Beyond the discrimination question, two structural findings from the topological representations survive scrutiny, each with a qualification.

#### Neighbourhood-based risk stratification

The four Mapper D^+^ features place each patient at a position in the graph built on the training cohort, and they are not redundant with one another. The largest pairwise correlation was |*r*| = 0.600 and the largest variance inflation factor was 3.318, both inside the prespecified thresholds. A patient can therefore be placed in a high-risk or high-success region of a precomputed atlas before any prediction is made. This supports a descriptive use, not a predictive one: three of the five lockbox configurations carry the D^+^ block and none improved discrimination by a meaningful margin. One qualification is essential: the D^+^ scores are derived from outcome-labelled node purities and are supervised summaries of the training cohort. Fold-wise construction prevented leakage, but their behaviour should not be read as topology discovering risk independently of the labels.

#### Variation in adverse-outcome composition across patient neighbourhoods

The Mapper graph comprised 99 nodes and 183 edges, with 11 nodes (11%) above the high-risk purity threshold and 72 nodes (73%) above the high-success threshold. Among the fifteen largest nodes, adverse-outcome purity ranged from 5.9% to 30.5%, a fivefold spread within a set of nodes that a single dominant-class label would treat as equivalent.

Whether this is clinically meaningful heterogeneity is a separate question. The stability ablation shows that the structure is reproducible under resampling and across a 3 × 3 parameter grid, with node-count CVs between 3.8% and 7.9% and purity standard deviations between 0.0098 and 0.0230, all inside prespecified thresholds. Reproducibility is necessary but not sufficient. The geometry is also metric-relative: Gower’s coefficient weights each variable equally [25], so the neighbourhood structure is a consequence of that encoding as much as of the underlying biology. Persistence summaries are provably stable to small perturbations of the point cloud [24], but that guarantee says nothing about the metric that defines the cloud.

### 4.3 Relation to prior work and evaluation recommendations

The evaluation problem generalises beyond this application. Studies applying topological methods to clinical prediction should report the following, alongside the items required by TRIPOD+AI [31]:

1. **The baseline, fully specified.** Report feature count, selection procedure and baseline performance before any comparison.
2. **Effect sizes with intervals, and a prespecified margin.** A null is informative only if the reader can see what it excludes.
3. **Stability of the representation as a primary result.** Sensitivity to lens, cover and clustering choices belongs in the main text, with thresholds fixed in advance.
4. **Outcome-informed and outcome-blind topological features reported separately.** They carry different evidential weight.
5. **One prespecified decision-analytic or calibration-based comparison**, such as decision curve analysis [30]. This distinguishes added structural information from movement along an unchanged ROC curve.
6. **The dissimilarity metric and the encoding of mixed-type variables.** Topological output is not invariant to either.

Our structural findings agree with the established use of Mapper for subgroup discovery. Nicolau et al. identified a breast cancer subgroup with a distinct survival profile that conventional stratification missed [8], and later work has recovered homogeneous patient subgroups in clinical data by similar means [51]. Those studies claim subgroup discovery, not incremental discrimination over a tuned baseline, so our null addresses a different claim and does not contradict them.

Our result diverges from studies reporting discriminative gains. The most plausible explanation is the input representation. Xin et al. [52] achieve an AUC of 0.767 and 0.875 from two clinical classification tasks for Multiple Sclerosis by applying persistent homology directly to inter-lesion spatial structure in MRI, a setting where topology is an intrinsic property of the image rather than a derived summary. Somasundaram et al. [53] show that cubical persistent homology features from lung CT scans associate with survival independently of tumour size, again extracting signal from unstructured pixel-level data. Our input was a tabular schema in which geometrically relevant structure had already been extracted by feature engineering.

These comparisons suggest that topological augmentation may contribute most when the input representation is unstructured and least when it has already been curated by feature engineering. This is a hypothesis for future work rather than an established general principle. The broader pattern is familiar. A systematic review found no performance benefit of machine learning over logistic regression for clinical prediction [5]. Our result is the same finding one level down, at the representation rather than the algorithm. Multimodal and radiological data carry real prognostic signal in tuberculosis [54]; our result suggests that a well-specified tabular schema already captures much of it.

### 4.4 Limitations

The cohort came from a single surveillance programme. It spans multiple countries, but referral, imaging and reporting practices within one programme are correlated, and no external cohort was available. Generalisability to routine programmatic settings is untested. We collapsed longitudinal records to one row per patient by worst-case aggregation, so every binary indicator became an “ever recorded” flag. In particular, Lost to follow-up was coded as an adverse (Negative) outcome alongside Died and Failure. Loss to follow-up is often a surveillance failure rather than a direct proxy for poor clinical outcome, and sensitivity to this coding choice was not tested. More broadly, the worst-case aggregation uses findings recorded across the full follow-up period rather than information available at a defined baseline time point. The resulting model reflects the complete clinical trajectory and cannot be directly applied to early treatment decisions without a prospective baseline cohort.

Gower’s coefficient, the UMAP lens and DBSCAN clustering were each reasonable choices not compared against alternatives. The null is bounded by the intervals reported in Section 3.4 rather than by a designed detectable effect. A larger or more heterogeneous cohort, or a weaker baseline, could yield a different answer. The stability of the derived D^+^ feature values themselves under Mapper parameter variation is a separate, open question not addressed by the graph-level ablation reported in Section 2.6.1.

## 5 Conclusions

Topological structure contributed descriptively rather than discriminatively to this clinical prediction task. Mapper and local persistent homology provided a view of the patient manifold not captured by the clinical feature schema, but neither augmented AUROC over a well-specified 99-feature baseline — four of sixteen paired comparisons significantly favoured the baseline, and no upper confidence limit exceeded +0.014 AUROC units. The methodological contribution is an evaluation design in which every topological step is fitted inside the training partition and applied to sealed data, making the comparison interpretable, together with evidence that AUROC parity is compatible with a feature block carrying feature variation that is not linearly redundant with the clinical predictors under standard correlation diagnostics. The direct next step is to apply this design to cubical persistent homology on CT and chest radiograph images, where the input is unstructured and its geometric content has not already been extracted by feature engineering.

## List of abbreviations

*ANOVA*: Analysis of variance
*ATOL*: Automatic topologically-oriented learning
*AUC*: Area under the curve
*AUROC*: Area under the receiver operating characteristic curve
*DBSCAN*: Density-based spatial clustering of applications with noise
*DR-TB*: Drug-resistant tuberculosis
*FDR*: False discovery rate
*LR*: Logistic regression
*LTF*: Lost to follow-up
*MDR*: Multidrug-resistant
*NIAID*: National Institute of Allergy and Infectious Diseases
*PCA*: Principal component analysis
*PH*: Persistent homology
*PR-AUC*: Area under the precision–recall curve
*RBF*: Radial basis function
*RF*: Random forest
*ROC*: Receiver operating characteristic
*SHAP*: SHapley Additive exPlanations
*SMOTE*: Synthetic minority over-sampling technique
*SVM*: Support vector machine
*TB*: Tuberculosis
*TDA*: Topological data analysis
*TRIPOD+AI*: Transparent Reporting of a multivariable prediction model for Individual Prognosis Or Diagnosis, extended for artificial intelligence
*UMAP*: Uniform manifold approximation and projection
*WHO*: World Health Organization
*XDR*: Extensively drug-resistant
*XGB*: XGBoost (extreme gradient boosting)

## Supplementary information

There is no supplementary material.

## Declarations

### Ethics approval and consent to participate

This study analysed de-identified, publicly available surveillance records from the NIAID TB Portals programme. As no new data were collected and all records were previously anonymised by the data custodian, formal institutional ethics review was not required under the applicable regulations.

The authors obtained written consent from the NIAID TB Portals programme to use the dataset for academic research purposes only.

### Consent for publication

Not applicable.

### Availability of data and materials

The datasets analysed during the current study are available from the NIAID TB Portals programme through its official web portal (https://tbportals.niaid.nih.gov).

### Authors’ contributions

DTM conceptualised the study, curated the data, performed the formal analysis and wrote the original draft. MVV and FC conceptualised the study, supervised the work, and reviewed and edited the manuscript. All authors read and approved the final manuscript.

## Authors’ information

## Acknowledgements

We gratefully acknowledge the University of Johannesburg for providing research resources, including internet access, computing facilities and library services. We also thank the University of Johannesburg Accelerated Academic Mentorship Programme (AAMP) for supporting the dissemination of this research.

## AI and large language model (LLM) use

During the preparation of this manuscript, the authors used Claude (Anthropic, claude.ai) for language refinement, including assistance with converting doctoral thesis content into journal article format. The authors have reviewed and edited all AI-assisted content, take full responsibility for the accuracy and integrity of the published work, and confirm that no AI tool was used to generate, fabricate, or alter any data, results, figures, or scientific conclusions reported in this study.

